# Reducing motion artefact in high resolution 7T MRI using the Magnetic Resonance Minimal Motion (‘MR-MinMo’) head stabilisation device

**DOI:** 10.1101/2025.10.10.25337727

**Authors:** Jyoti Mangal, Simon Richardson, Yannick Brackenier, Matthew Gardner, Pierluigi Di Cio, Chiara Casella, Shaihan Malik, Jo Hajnal, Martina F Callaghan, Fred Dick, David W Carmichael

## Abstract

**Purpose:** To evaluate the effectiveness of the MR-MinMo head stabilisation device in mitigating motion artifacts during long-duration, high-resolution 7T MRI scans, with and without retrospective motion correction.

**Methods:** The MR-MinMo was tested on 7 paediatric and 12 adult healthy volunteers using 0.6mm isotropic, 3D Multi-Echo Gradient Echo (ME-GRE) scans. A ∼10-minute ME-GRE scan (linear sampling acceleration factor 2×2) and a ∼20-minute scan (DISORDER sampling acceleration factor 1.4×1.4) were obtained. Both scans were acquired with and without the MR-MinMo in a 2×2 factorial study design. Qualitative and quantitative assessment of image quality of the first echo image was obtained via visual inspection and the normalized gradient squared (NGS) metric respectively. A repeated measures ANOVA was used to determine individual and combined effects of the device and DISORDER images with retrospective motion correction on NGS values, with age as a binary factor (paediatric/adult). T2* maps were generated and the standard deviation of white matter R2*(=1/T2*) values assessed across conditions.

**Results:** The MR-MinMo significantly reduced motion artifacts both visually and in terms of NGS scores, particularly in paediatric volunteers. There was a significant interaction between MR-MinMo and DISORDER motion correction, suggesting that MR-MinMo improved retrospective motion correction. T2* maps demonstrated improved visual appearance and reduced WM variance with the MR-MinMo.

**Conclusion:** The MR-MinMo can improve image quality via motion reduction in high-resolution 7T scans By keeping motion within a correctable regime, the device can also improve the performance of retrospective motion correction methods.

## Introduction

Among imaging modalities, MRI is particularly sensitive to subject motion compared to alternatives like ultrasound or CT. This heightened sensitivity arises from the longer acquisition times required for most MRI sequences to gather sufficient data for faithful image formation. Since acquisition times are far longer than timescales of physiological, volitional and accidental subject motion, they often affect MRI image quality^1,2^. When motion occurs at a spatial scale comparable to the imaging resolution, it introduces artefacts including reduced signal-to-noise ratio (SNR), distortion, blurring, and ghosting. Even in compliant volunteers, degradation may be caused by periodic involuntary motion e.g. cardiac, respiratory and blood flow, aperiodic motion including yawning, swallowing, sneezing or coughing, and conscious movement e.g. due to discomfort^3^. In children and in patients with neurological disorders motion tends to be exacerbated^1,4^.

Ultra-high field MR (≥7T) offers the advantage of increased SNR, enabling the acquisition of high-resolution images. However, their acquisition requires longer scan times, making them even more susceptible to motion-induced degradation. Reduced voxel dimensions also increase sensitivity to finer scale subject motion. Strong interactions at ultra-high fields may pose additional challenges - motion-induced static magnetic (B₀) field changes^5^ increase due to greater susceptibility effects, while applied Radio Frequency (B₁) fields also change related to movement^6^ due to stronger coupling between the subject and RF field, increasing the complexity of motion correction strategies.

Quantitative mapping such as R2* mapping is sensitive at 7T and enables tissue characterisation of varying iron content^7,8^ with increased specificity^7^. This may be important in several neurological conditions including Parkinson’s disease and epilepsy^8^. However, motion can severely degrade the precision and accuracy of the R2* estimates^9–11^.

To fully realise the image quality benefits of 7T MRI for clinical usage, it is crucial to develop practical solutions for reducing motion effects appropriate for paediatric and adult neuroimaging. To reduce motion at source by providing improved head stability and comfort, we designed and tested a new head stabilization device for high resolution 7T neuroimaging, named *MR-MinMo*^12^. A particular aim was to investigate if it improves retrospective motion correction. To test this, we used a self-navigated sequence approach that does not require any additional hardware, namely Distributed and Incoherent Sample Orders for Reconstruction Deblurring using Encoding Redundancy (DISORDER)^13^. This sequence involves modified sampling of a cartesian k-space trajectory with an iterative retrospective reconstruction that includes a motion estimate. We hypothesized that the MR-MinMo device would improve image quality for DISORDER retrospective motion correction by maintaining the data within a correctable regime.

## Methods

### The MR-MinMo Device

The MR-MinMo device used in this study is a head stabilization tool designed to reduce motion in awake participants, typically aged 6 and older. The device version used in this study was a prototype for comfortable head positioning inside the 8 and single channel transmission 7T RF head coils manufactured by Nova Medical (Wilmington, MA, USA).

In brief, the MR-MinMo consists of a polycarbonate frame which conforms to the inner surface of the Nova coil and serves as mounting points for various functional modules. These modules are inflatables and fabric covered pads which are at set positions. The frame includes an articulated ‘halo’ which hinges down, and latches closed to allow entry of the participant and rapid egress. The diagram of the device in its open and closed configuration is shown in Figure 1. The halo’s latches can be unlocked and locked to change the device’s configuration. When locked, the halo is latched to the lower frame in ‘closed configuration’ and ready for scanning. When unlocked the halo can be lifted for the device to be in ‘open configuration’ and ready for loading/unloading the participant. Additionally, the use of a hairnet with the MR-MinMo is recommended to distribute the gripping force on the head, to prevent hair being caught in the mechanism and to provide a more feel compared to the plastic surface of the inflatables.

**Figure 1.**
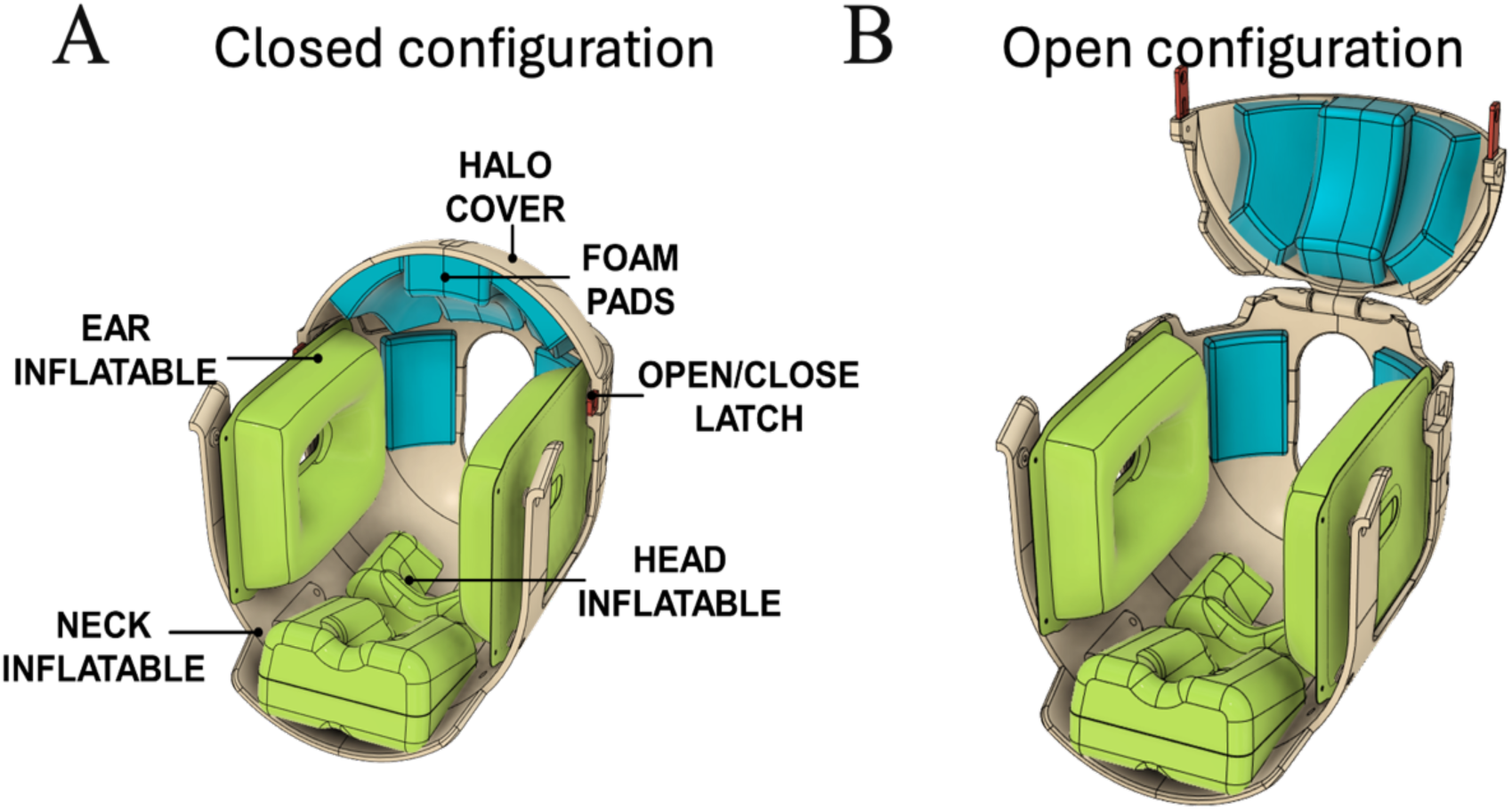
The schematic diagram of the MR-MinMo with labelled components in (A) closed configuration and (B) open configuration. The device primarily consists of (i) Frame: the plastic structure that holds the other components in place within the head coil (ii) Halo: the upper unhinged section of the main Frame, (iii) Latch: the lock that holds the unhinged section of the main frame, (iv) Inflatables: the various plastic components that can be inflated or deflated around the ears, neck and head, (v) Padding system: consists of horn pad and mohawk pad made with memory foam, and (not shown) (vi) Manifold: the box that controls the air pressure distribution specific to each inflatable and (v) Vent: the valve mounted on the manifold which releases air from all inflatables at once when set to Open.

The MR-MinMo has been designed with a quick release valve on the manifold to facilitate quick subject evacuation and also includes a relief valve to assure pressure does not exceed safe levels. Most participants are able to have a clear line of sight out of the coil via a mirror. This enables standard patient distraction and facilitates functional imaging task-based studies.

### Image Acquisition

To test the effectiveness of the MR-MinMo device, we utilized a high-resolution 3D ME-GRE acquisition. Two types of k-space sampling were used: 1), a standard (or conventional) linearly encoded acquisition; and 2), a DISORDER encoded acquisition. The ME-GRE acquisition was chosen because it facilitates T2* estimation, which in turn is sensitive to both bulk head and physiological motion. The DISORDER trajectory served two purposes. First, it introduced a deliberately motion-sensitive trajectory that is likely to produce artefacts even in compliant volunteers prior to motion correction, making it a more sensitive test of the MR-MinMo device’s capacity for reducing small-scale motion. Second, it provided data that could be used for retrospective correction, thereby allowing for comparison of efficacy and potential synergistic application with the MR-MinMo device.

The parameters used as a base for image acquisition paradigms were TR=30ms, flip angle = 36°, bandwidth = 470Hz/px, number of echoes = 10, equally spaced at 2.68ms intervals between TE1 = 2.27 ms and TE10 = 26.39 ms. Bipolar readouts were acquired with the elliptical shutter switched on for scan acceleration. A non-selective RF pulse was used, and echo readouts were optimised to minimize dead time. The scan had an isotropic resolution of 0.6mm³, and a field of view (FOV) = AP / LR / HF = 256 x 173 x 218mm³.

In the DISORDER encoding method (Figure 2), k-space samples are acquired in a pseudo-random order from rectangular regions or ‘tiles’ of the phase encoding plane of k-space. Acquisition is performed in shots, where each shot consists of acquiring one sample from each tile within the phase-encoding plane *k*_2_x *k*_3_and ‘shot duration’ is defined as the time taken to acquire each shot. In Figure 2, each colour represents a different shot. As shown in Cordero-Grande et al^13^, each sample from within a tile is acquired pseudo-randomly resulting in a distributed temporal coverage within each tile as well as for the whole k-space spectrum. In this study, we employed the ‘random-checkered’ approach, with data acquired in the head-foot (HF) *k*_1_, anteroposterior (AP) *k*_2_and left-right (LR) *k*_3_ orientations. Rotations of the sagittal plane are sampled faster within each shot, improving robustness to intra-shot motion.

**Figure 2.**
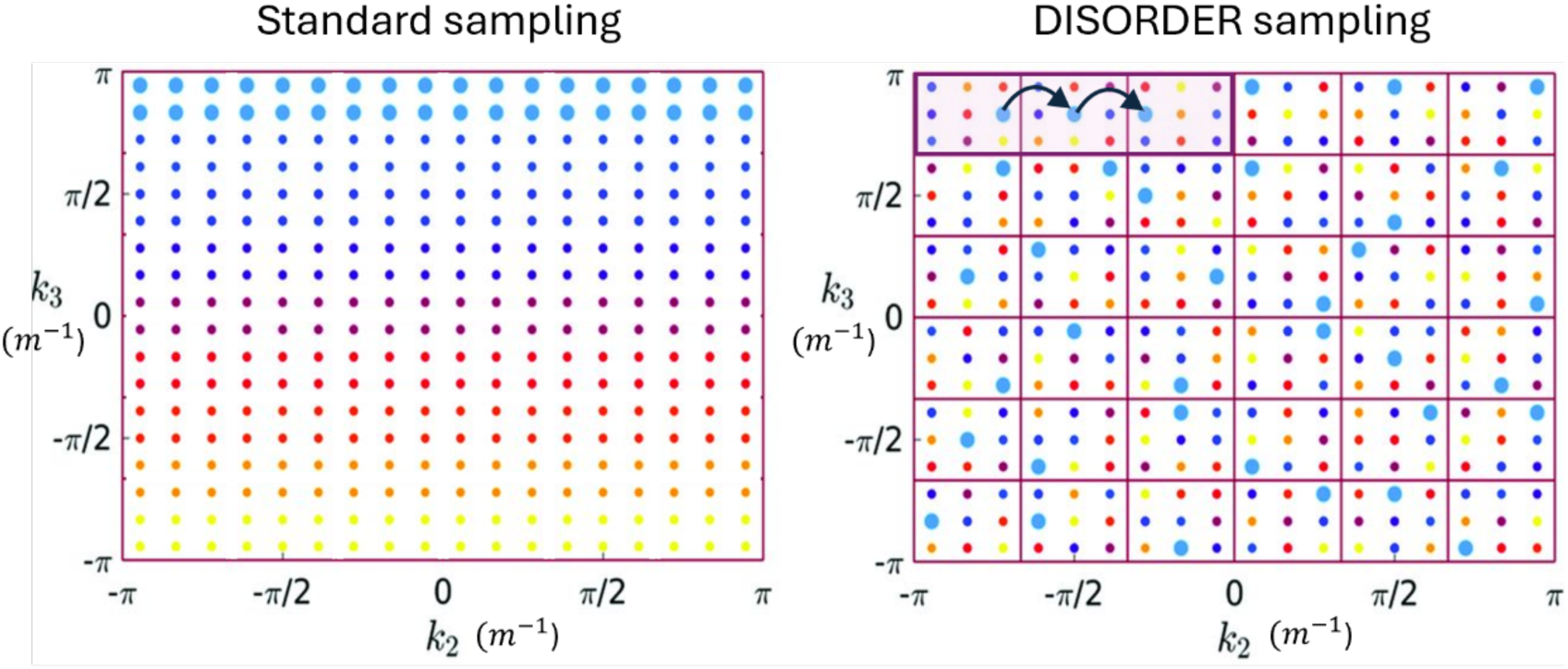
The different k-space data acquisitions shown for visual representation. On the left, the standard sampling case in which adjacent lines in the phase encoding plane are acquired linearly. On the right, the DISORDER sampling case is shown for a tile size of 3×3 in which distributed information in k-space is acquired with a pseudo-random order of samples. Each colour represents a different shot.

Informed consent was obtained from all participants or their legal representatives, as appropriate. All healthy adult scans were performed according to the local ethics approval (HR-18-19-8700).

#### DISORDER scan parameters

Previous work by Cordero-Grande et al^13^ demonstrated robust motion correction with DISORDER when the acceleration factor was at most 1.4. This was replicated here on the 7T Siemens Terra scanner that only allows for integer acceleration factors by oversampling in the phase encoding (PE) directions. To achieve the target acceleration factor of 1.4×1.4, a GRAPPA^14^ acceleration of 2 was used in both PE directions, together with oversampling by 44% and 42% in each phase encoding direction respectively. Oversampling percentages ‘P’ were determined using the scanner-permissible values, calculated according to the formula: 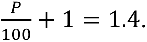 The tile size for the DISORDER acquisition was set to 24 x 24 in the PE plane, implying a total of 576 points in each tile as well as 576 total shots or motion states in the k-space, with a shot duration of 2.21s. This tile size was selected within the scanner’s allowable limits while ensuring quicker shots to optimise motion correction. Motion was estimated from k-space data segments of 6 consecutive shots. This was done to balance the timescale of motion states while remaining within practical computational times for 0.6mm^3^ resolution images. The total acquisition time for the DISORDER encoded scan was 21:01min, classified as a “very long scan”.

#### Linear sampling scan parameters

A high-resolution scan was performed using the standard linear Cartesian sampling trajectory. This acquisition was designed to be faster and less sensitive to motion compared to the scan using DISORDER sampling. To achieve this, the acceleration factor was set to 2×2 with no oversampling in the phase encoding directions, providing an overall acceleration factor of 4. The total acquisition time for this scan was 10:38min and while it was shorter than the DISORDER scan, it was classified as a “long scan”. This acquisition was chosen as typical for conventional ME-GRE studies used for quantitative mapping^15^.

To estimate coil sensitivity maps for offline reconstruction of the high-resolution scans, a fully sampled reference GRE scan was acquired using the same field of view (FOV = 256 x 173 x 218mm³) and adjustment volume. No oversampling was applied, and the elliptical shutter was switched on. The acquisition parameters were set to flip angle = 18°, TR = 5.3ms, with a single echo readout at TE = 2.27ms and bandwidth = 420Hz/px. The resolution for the reference scan was 4.7mm³, with a total acquisition time of 14s. As for the high-resolution scans, a non-selective RF pulse was used. The estimation of receiver coil sensitivity maps needed for the implementation of the reconstructions used the ESPIRiT algorithm.

### Image Reconstruction

For all images, reconstruction was performed off-line using code developed by the authors in MATLAB version R2019b (www.themathworks.com). All images were initially reconstructed using conjugate gradient SENSE (CG-SENSE). The CG-SENSE reconstruction was performed with a maximum of 300 iterations, with the tolerance level set to 0.001.

For DISORDER image reconstruction with motion correction, a least squares minimization problem known as the aligned-SENSE^16^ was solved as previously described^13^. In this approach, initialization is a CG-SENSE reconstruction. Rigid body motion states, as well as the image data are alternately minimized. Motion estimates are facilitated by the multiple views and images acquired from different coils. These views provide complementary information that helps constrain the motion estimation problem, enabling it to be solved iteratively.

For the motion estimation, the reconstruction framework used two resolution levels, defined as the pyramid plan by Cordero-Grande et al^13^. These levels corresponded to downscaled resolutions at factors [0.5, 1] of the total resolution resulting in voxel sizes of [1.2, 0.6mm]. Motion parameters were estimated using 4 iterations at the first resolution level (1.2mm voxel size). The final reconstruction was then performed at the higher resolution level (0.6mm voxel size) using the motion parameters estimated from the first resolution level, with only 1 iteration performed at this resolution to maximize computational efficiency^13^. In addition to motion estimation, an outlier rejection step was incorporated to enhance the quality of the reconstruction. Outliers were identified based on the normalized median error for each motion state, enabling the pipeline to ignore data points likely affected by excessive motion. The total reconstruction pipeline, in its current version, for the DISORDER dataset took approximately 8-10 hours for level 1 and 20-30 hours for level 2 depending on the motion correction pipeline. Reconstructions were performed on a dual (8×2 cores) Intel(R) Xeon(R) @3.10Gz system with 256GB RAM.

All image volumes were corrected for bias field inhomogeneities using SPM12’s bias correction algorithm^17,18^, processed via the ‘Segment’ batch.

### 2×2 Factorial Study Design

A 2×2 factorial design was used (see Figure 3) to assess the independent and interactive effects of using the MR-MinMo device and DISORDER sampling with retrospective motion corrected reconstruction with the following four conditions: A) Standard padding with linear sampling, B) MR-MinMo with linear sampling, C) standard padding with DISORDER sampling and motion correction and D): MR-MinMo with DISORDER sampling and motion correction. This allows the assessment of the effect of MR-MinMo compared to standard padding and the potential additional benefit to reducing motion with MR-MinMo at source prior to retrospective correction. Across all the healthy volunteers scanned, the order of using the MR-MinMo and standard padding was randomized to counterbalance any potential order effects, as well as the known interaction between duration of time in the scanner and participant motion^19^.

**Figure 3.**
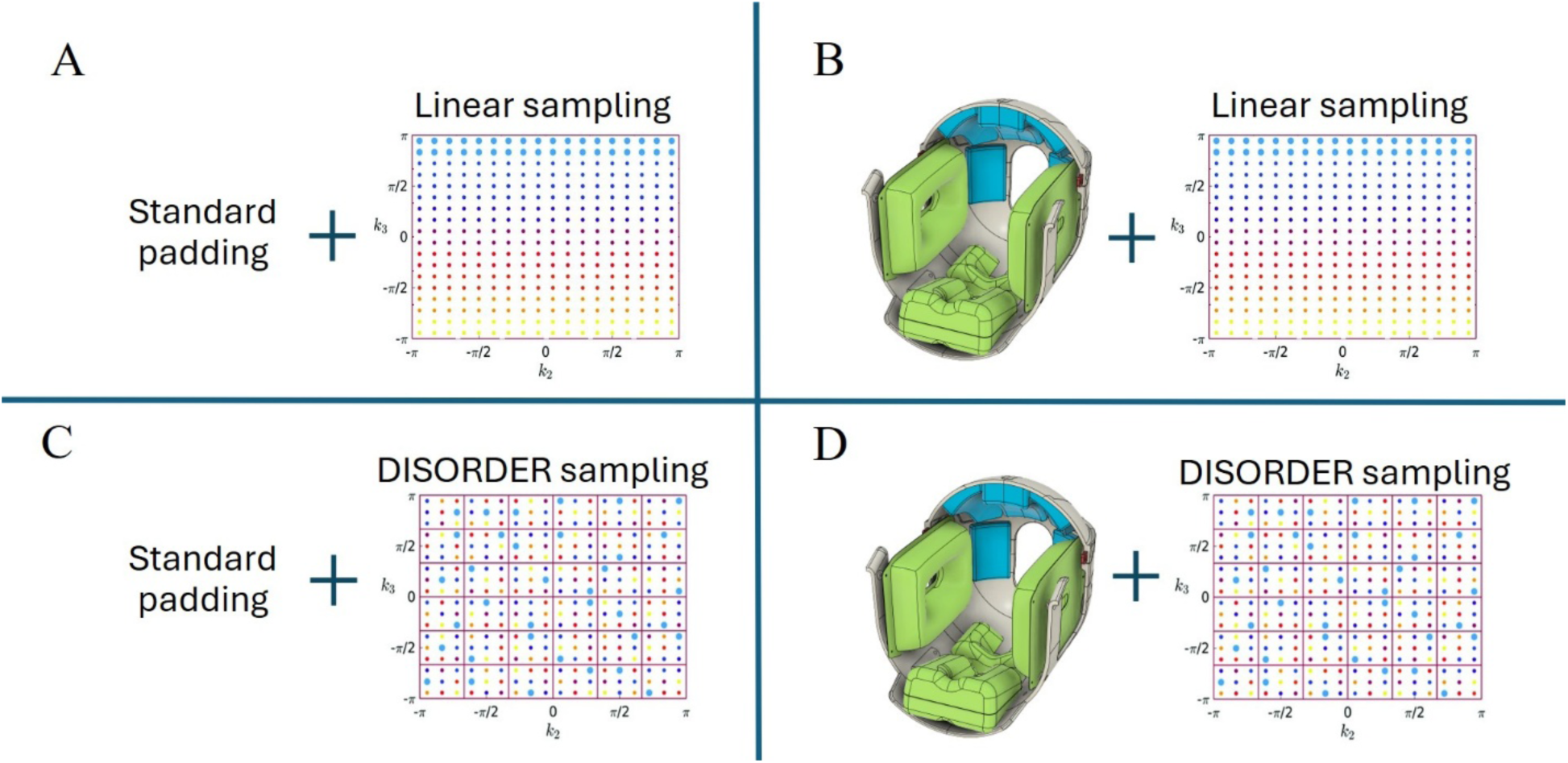
The 2×2 experimental scanning scheme. The independent variables of the scheme were MR-MinMo and DISORDER with two levels of each i.e. MR-MinMo, standard padding and DISORDER, linear sampling, resulting in 4 conditions: A) Standard padding + linear sampling; B) MR-MinMo + linear sampling; C) standard padding + DISORDER; and C) MR-MinMo + DISORDER. Standard padding implies the conventional padding set-up was used. DISORDER referred here implies DISORDER trajectory and motion correction was used.

### MR-MinMo device set-up

The MR-MinMo device was used during scan acquisition in the closed configuration as shown in Figure 1(A). While loading the participant with the device, constant feedback was obtained by the radiographer on participant comfort and stability. The hairnet size used by each participant was chosen with individual verbal feedback in the radiographer’s presence to ensure the correctly sized hairnet was used.

Before loading the participant, the device was seated in the head coil in open configuration and was stabilized purely by a friction fit. The participant was asked to lie on the table while being guided to position their head in the device. Once the head was positioned into the device, the participant was instructed to shuffle the head down into the coil to avoid any empty spaces near the scanner-end of the coil. Minimal air was pumped into the neck inflatable by squeezing the bulb twice lightly to give neck support while the participant was instructed to raise their chin slightly. The halo was then secured in the closed configuration, and the neck, ear and head inflatables were further inflated with air until the participant reported experiencing immobilization while still maintaining high comfort levels. While using the MR-MinMo, no additional foam or cushioning was used.

The total time that the MR-MinMo was used during a single scan session was ∼33 mins: a DISORDER acquisition (∼21 mins), a standard acquisition (∼11 mins), and a combination of reference acquisition and localizer (∼1 min). This was roughly half of the total scan session time, with the other half performed without the MR-MinMo using the standard participant set-up and padding used at our institution and the same image acquisition protocols. (Again, the order of MR-MinMo and standard scan was pseudorandomized across participants). Each scan session lasted ∼1 hour and 15 minutes, with approximately 5 to 10 minutes to reposition the participant.

Participants were positioned by experienced research radiographers with and without the MR-MinMo device. The standard padding involved the use of a manufacturer-provided foam pad being placed under the head, and pads filling the space between the ears and the outside of the coil. For both setups, a cushion was placed under the knees for comfort and stabilization as conventionally used, and earbuds provided for sound attenuation.

To minimize discomfort or boredom, all the paediatric participants watched TV (without sound) for the entire session via a projector external to the scan room and screen mounted within the scanner bore. Adult participants, many of whom were accustomed to MRI, were offered TV although not all chose to watch.

### Participants

20 participants were recruited for the study (12 adults and 8 children). All participants were above the minimum weight requirement of 30 kg for the scanner. Out of the 20, 19 participants completed the scanning according to the 2×2 factorial design: 12 adults (ages 20–36 years, Mean 28.25; 7 females, 5 males) and 7 children (ages 10–15 years, Mean 12.14; 2 females, 5 males). The remaining 1 participant withdrew due to discomfort during the standard set-up condition. Data from the linear Cartesian encoded acquisition was successfully processed for all 19 participants. For the DISORDER encoded acquisition, data from 16 participants (9 adults and 7 children) were processed, with 3 adult datasets being unavailable due to technical raw data transfer issues. The paediatric participants were healthy volunteers (HVs) experiencing both 7T MRI and the MR-MinMo device for the first time. In contrast, most adult HVs were familiar with the research scanning environment and had previously undergone a 7T MRI.

### Image quality analyses

To assess the effects of both the MR-MinMo device and DISORDER trajectory on image quality, analyses were performed on all reconstructed ME-GRE image volumes (the shortest TE image, or echo 1, was used due to its highest SNR) and R2* maps. For the echo 1 image, the aim was to compare image quality across the different conditions in the 2×2 factorial design, using the normalized gradient squared (NGS) metric, which correlates closely with visual image quality assessment^20^. The NGS metric was calculated according to the formula following Ehman et al^20^

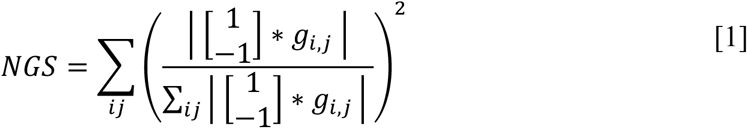

where *g_i,j_* is the pixel value at coordinate *i*, *j* in the image.

For image quality analysis, whole brain regions of interest (wbROIs) were extracted using SPM12. These were obtained using the scanner reconstructed first echo volume from the linear sampling and standard padding acquisition (condition A). This image was segmented and the tissue classes (grey matter, white matter, and CSF) were determined to be within the brain if they had a probability greater than 0.9 of belonging to one of these tissue classes. To ensure a complete brain ROI, MATLAB function *imclose* was used to fill any gaps, and *imerode* with a disk element of radius 2 was applied to remove boundary pixels. The within-wbROI mean NGS value was calculated for each of the four acquisitions resulting from the 2×2 factorial design.

### Motion parameters analysis

For each volunteer, motion differences between consecutive motion states (also termed frames) estimated during DISORDER motion correction reconstruction were calculated across the 96 motion states obtained. The resulting 95 element difference vectors per scan contained six parameters: three translational displacements (*Δx*, *Δy* and *Δz*) and three rotational displacements (*Δα*, *Δβ* and *Δγ*). The root mean square (RMS) displacement for each state transition was computed as

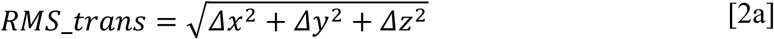

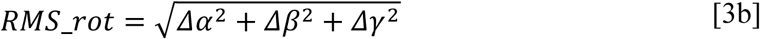

For each volunteer, the maximum and mean RMS_rot and RMS_trans values were determined by taking the mean and max value across all states for each individual for DISORDER scans with and without the MR-MinMo. Group statistics were calculated by averaging these individual maximum and mean values for each condition (MR-MinMo and standard padding) on a per participant basis.

### T2* map analysis

For T2* precision assessment using linear sampled scans, the motion degradation index SD(R2*) was calculated following the validated metric from Castella et al^11^. This index quantifies motion artefacts by measuring the standard deviation of R2* (R2* = 1/T2*) values within white matter, where larger values indicate greater motion degradation. White matter was segmented using the same tissue classification approach on first echo images, with probability maps thresholded at 0.9 and processed using the same morphological closing (disk element radius 2). T2* maps were converted to R2*, outliers (values outside 5-200 s⁻¹) were removed, and SD(R2*) was computed within each white matter mask. Lower SD(R2*) values indicate improved precision attributable to reduced motion artifacts.

## Results

### MR-MinMo efficacy

MR-MinMo images were visually improved in nearly all subjects both for the long duration linearly sampled acquisition and for the very long duration DISORDER retrospective motion correction images. This can be seen in Figure 4 and Figure 5 that show the bias field corrected first echo image of the 9 adults and 7 children, for the MR-MinMo (B & D) and standard padding conditions (A & C). The images show the zoomed-in regions near the frontal cortex for all participants where motion can be most clearly visualized. While motion artefacts were visually reduced for nearly all participants when the MR-MinMo device was used, the improvement in image quality was more apparent in paediatric HVs. Since the position within the RF coil of the HV had changed between the MR-MinMo and standard padded scans, slices with the closest visual appearance without any explicit co-registration are shown to avoid interpolation effects.

**Figure 4.**
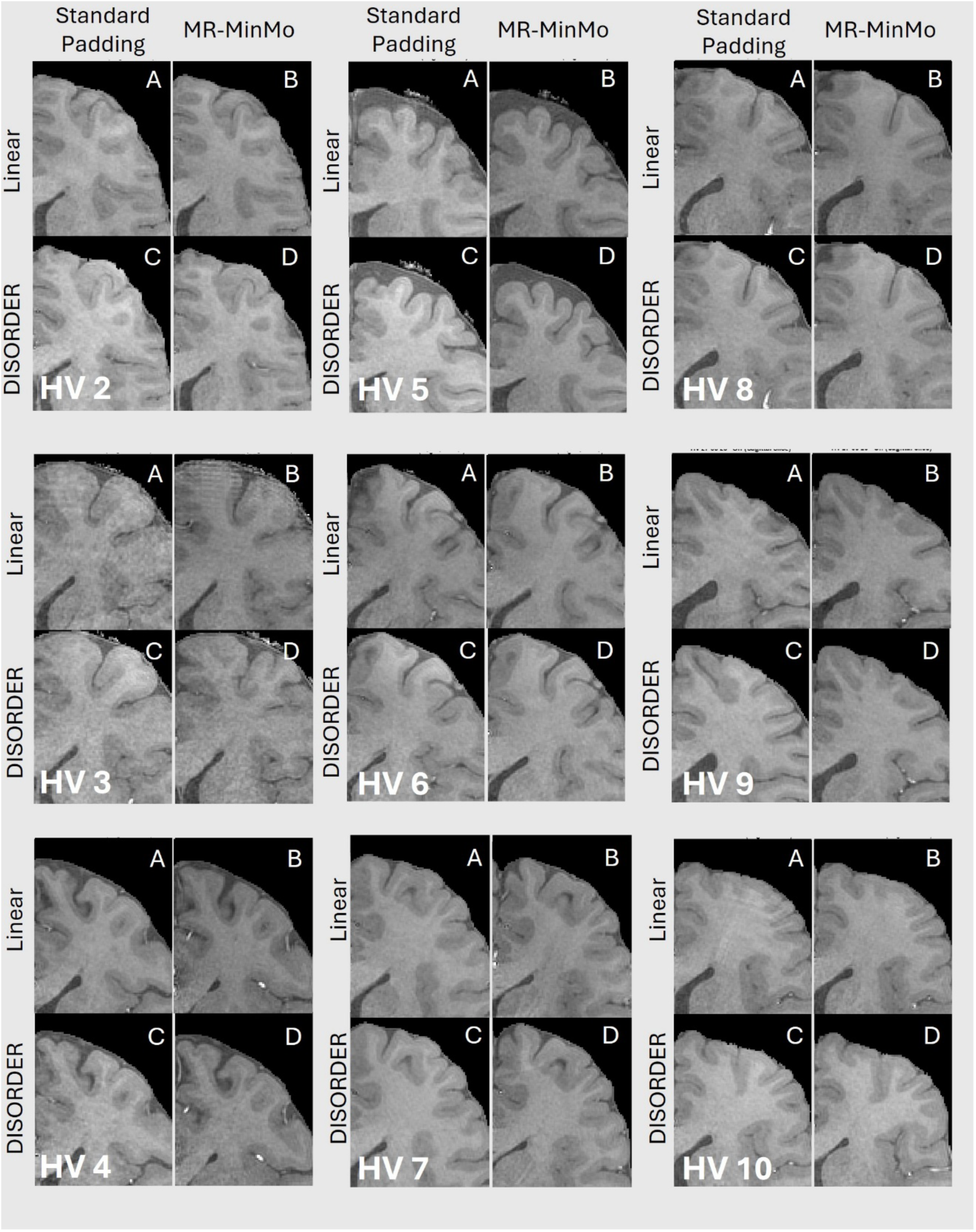
Representative images showing the standard padding (SP) and MR-MinMo conditions for the linear sampling and retrospectively motion corrected DISORDER sampled scans for 9 adults HVs (HV2-HV10). Images shown are zoomed regions of transverse orientation slices to facilitate visualisation of motion artefact levels. The images were selected visually to display the closest equivalent anatomical area without performing coregistration between SP and MR-MinMo conditions that required subject repositioning.

**Figure 5.**
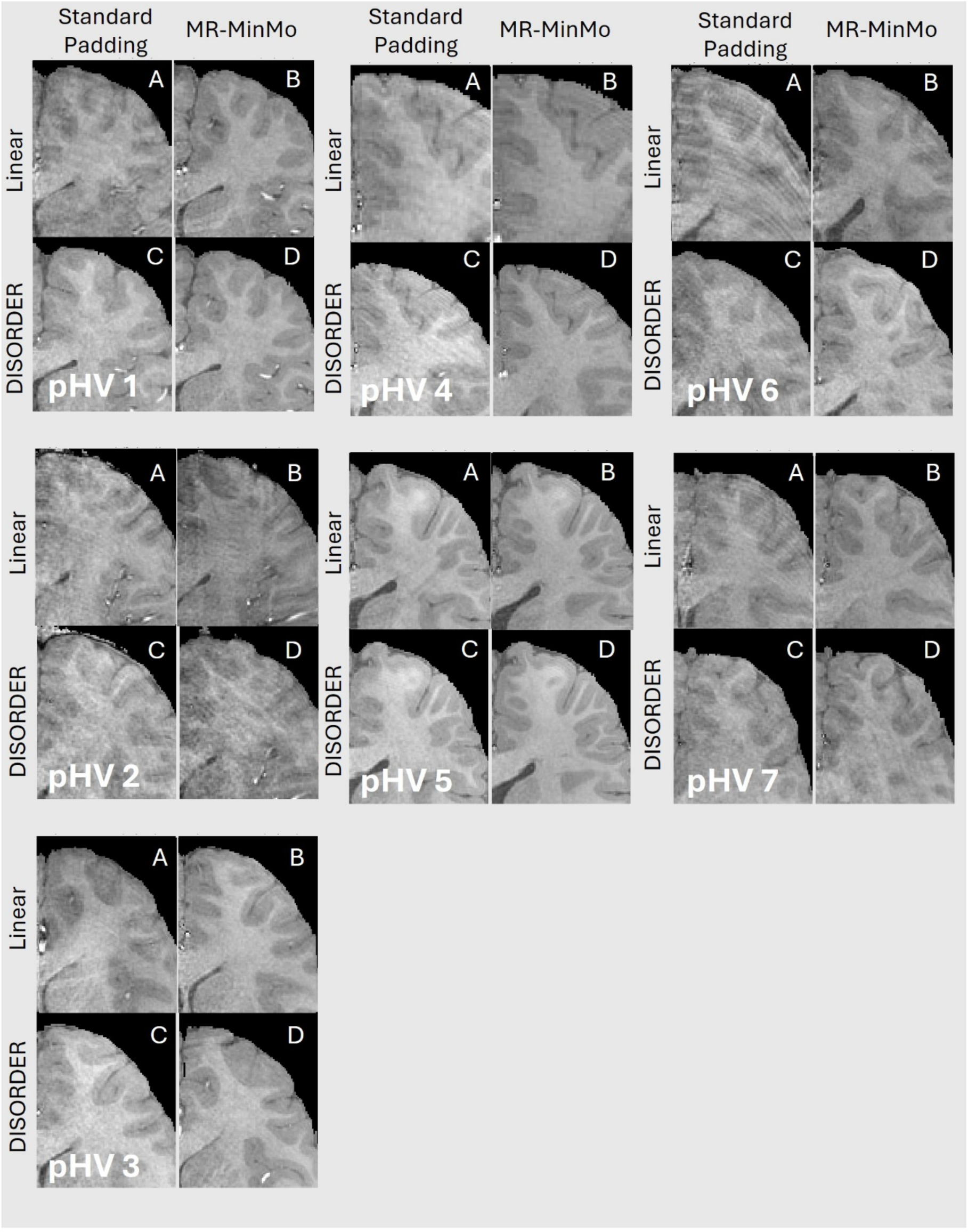
Representative images showing the standard padding (SP) and MR-MinMo conditions for the linear sampling and retrospectively motion corrected DISORDER sampled scans for 7 paediatric HVs (pHV1-7). Images shown are zoomed regions of transverse orientation slices to facilitate visualisation of motion artefact levels. The images were selected visually to display the closest equivalent anatomical area without performing coregistration between SP and MR-MinMo conditions that required subject repositioning.

### Quantitative image quality analyses

To quantify the artefact levels in the ME-GRE images shown above, the whole brain NGS values were calculated for all the HVs and experimental conditions and are shown graphically in Figure 6. Figure 6A & Figure 6B shows the NGS values calculated from the images acquired with linear sampling and DISORDER motion correction respectively. Each plot shows every individual’s NGS values paired between standard pads and MR-MinMo. It can be seen that the NGS values are increased, on average and for most individuals for both linear and DISORDER scans. Figure 6(C) show the NGS values calculated from the images acquired with standard padding and the DISORDER trajectory prior to and post retrospective motion correction. This demonstrates that the reconstruction was effective in reducing motion in the DISORDER acquired images. Finally, in Figure 6(D) the NGS values were calculated prior to and post DISORDER-based retrospective motion correction of images acquired with the MR-MinMo. Comparing between Figure 6C and Figure 6D it can be seen that the MR-MinMo NGS values (Figure 6D) are increased compared to the standard pads and that the combination of MR-MinMo and correction provides an increased NGS value compared to the retrospective correction or the MR-MinMo used individually. These differences were statistically significant; the two-way repeated measures ANOVA with age as a binary factor showed that there was a significant main effect of MR-MinMo on the NGS values (F=34.393, p<0.001). A significant interaction was found between MR-MinMo and age group (F=8.311, p=0.012), and a significant three-way interaction between MR-MinMo, DISORDER and age group (F=5.599, p = 0.033) indicating differential effects of the MR-MinMo device across age groups and sampling types. Overall, this implies that DISORDER was more effective when the range of motion was reduced by the MR-MinMo device. DISORDER motion correction efficacy was assessed by comparing NGS values before and after retrospective correction using paired t-tests. As expected, retrospective motion correction significantly improved DISORDER scans across subject age groups and head stabilisation conditions: adult HVs standard padding (p=0.0016), adult HVs MR-MinMo (p=0.0002), pHVs standard padding (p<0.0001), and pHVs MR-MinMo (p=0.0012).

**Figure 6.**
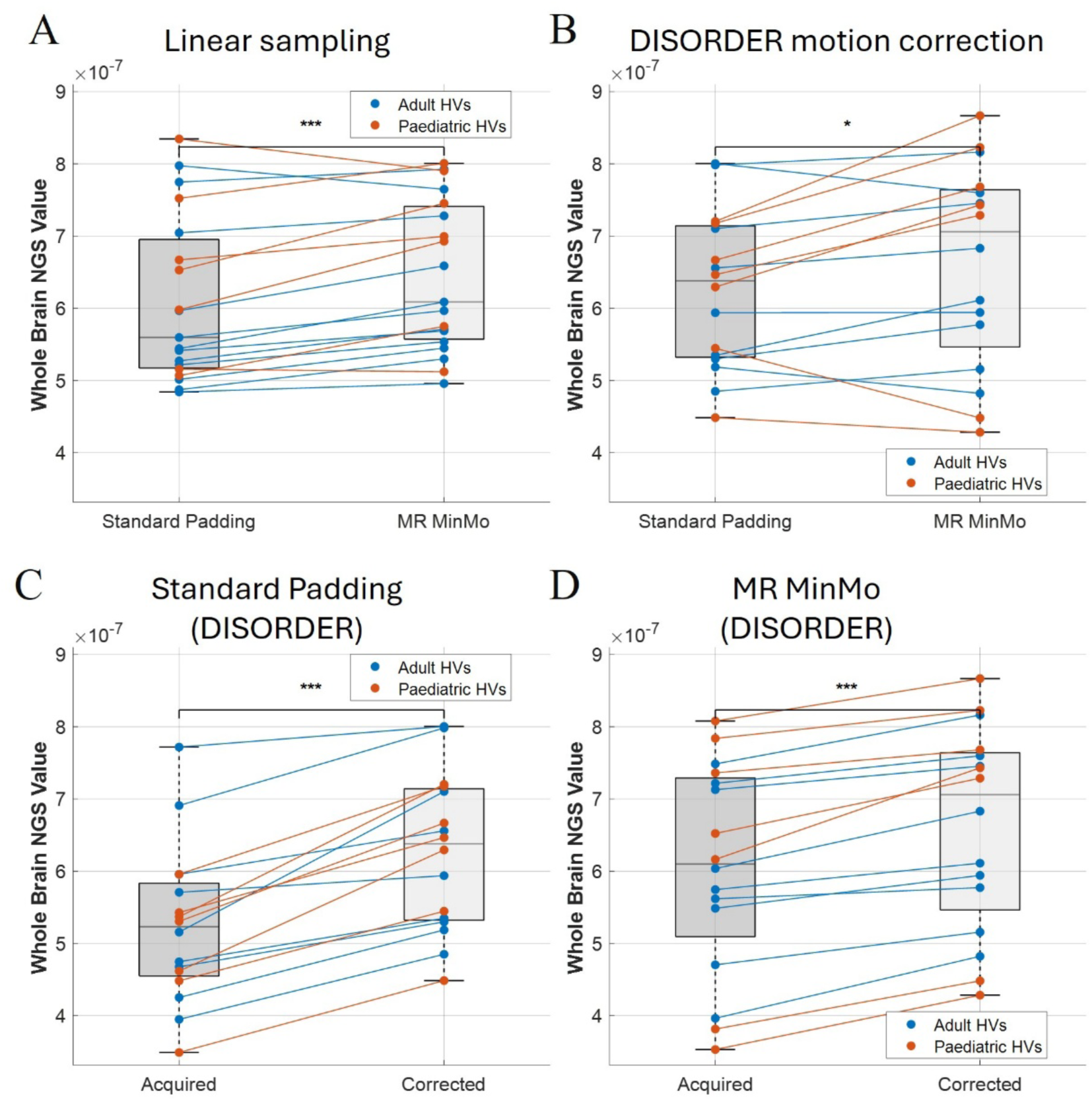
Boxplots of the whole-brain NGS values for all participants, comparing the SP and MinMo conditions in (A) which shows the results for the linear sampling acquisition and (B) for the motion corrected DISORDER case,; Also comparing the DISORDER acquired and DISORDER motion corrected cases (C) for the standard padding case and the D) for the MR-MinMo case. The connected lines represent the change in image quality between the different conditions for adult (blue) and paediatric HVs (red) separately, with an increase in NGS indicating improved image quality. The boxplots display the median NGS value, while the box boundaries represent the 1st and 3rd quartile range. Asterisks denote statistical significance (*p < 0.05, ***p < 0.001)

### Motion quantification

As calculated from the motion states estimated by DISORDER, MR-MinMo provided substantial motion reduction in both volunteer groups, with greater effects observed in paediatric participants (**Table 1**). Overall, MR-MinMo reduced mean framewise translation by 36% (from 0.17 to 0.11 mm) and mean rotation by 37.0% (from 0.091° to 0.057°). Paediatric HVs showed larger absolute motion reductions (mean translation: 41% reduction from 0.263 to 0.156 mm; mean rotation: 43% reduction from 0.153° to 0.087°) compared to adult HVs (mean translation: 26% reduction from 0.096 to 0.071 mm; mean rotation: 22% reduction from 0.044° to 0.035°). Maximum motion parameters showed similar patterns, with overall reductions of 37% for translation and 26% for rotation. These measurements indicate that paediatric participants moved substantially more than adults during the DISORDER sampled acquisition, but both groups benefited substantially from MR-MinMo.

**Table 1.** Framewise head pose values across HVs for translation and rotation are presented for standard padding (SP) and MR-MinMo (MR-M) conditions, with percentage reduction (Red%) indicating the motion reduction achieved with the MR-MinMo device. Individual volunteer maximum and mean framewise rotations and translations were averaged (represented by <max> and <mean>) within each group (Adult, Paediatric) and across the combined group (All) for the group statistics. A frame represents a motion state estimated from the DISORDER retrospective correction framework with a temporal resolution of ∼13s used here.

|  | <Mean> Rotation (°) |  |  | <Max> Rotation (°) |  |  |
| --- | --- | --- | --- | --- | --- | --- |
|  | SP | MR-M | Red% | SP | MR-M | Red% |
| Adult | 0.044 | 0.035 | 21.8 | 0.213 | 0.153 | 28.2 |
| Paediatric | 0.153 | 0.087 | 42.8 | 0.622 | 0.469 | 24.6 |
| All | 0.091 | 0.057 | 37 | 0.388 | 0.289 | 25.6 |
|  | <Mean> Translation (mm) |  |  | <Max> Translation (mm) |  |  |
|  | SP | MR-M | Red% | SP | MR-M | Red% |
| Adult | 0.096 | 0.071 | 26.2 | 0.461 | 0.231 | 49.8 |
| Paediatric | 0.263 | 0.156 | 40.8 | 1.401 | 0.959 | 31.6 |
| All | 0.168 | 0.107 | 36.3 | 0.864 | 0.543 | 37.2 |

### T2* maps

Motion precision was quantified using the motion degradation index SD(R2*) within white matter across all volunteers. MR-MinMo demonstrated significantly improved T2* maps compared to standard padding (as shown in Figure 7A), with mean SD(R2*) values across the group of 12.0 s⁻¹ for MR-MinMo versus 13.1 s⁻¹ for standard padding (p < 0.05, paired t-test). The significantly lower standard deviation with MR-MinMo (12.0 vs 13.1 s⁻¹) indicated more consistent results across all volunteers, demonstrating reduced motion-related variability in R2* measurements.

**Figure 7.**
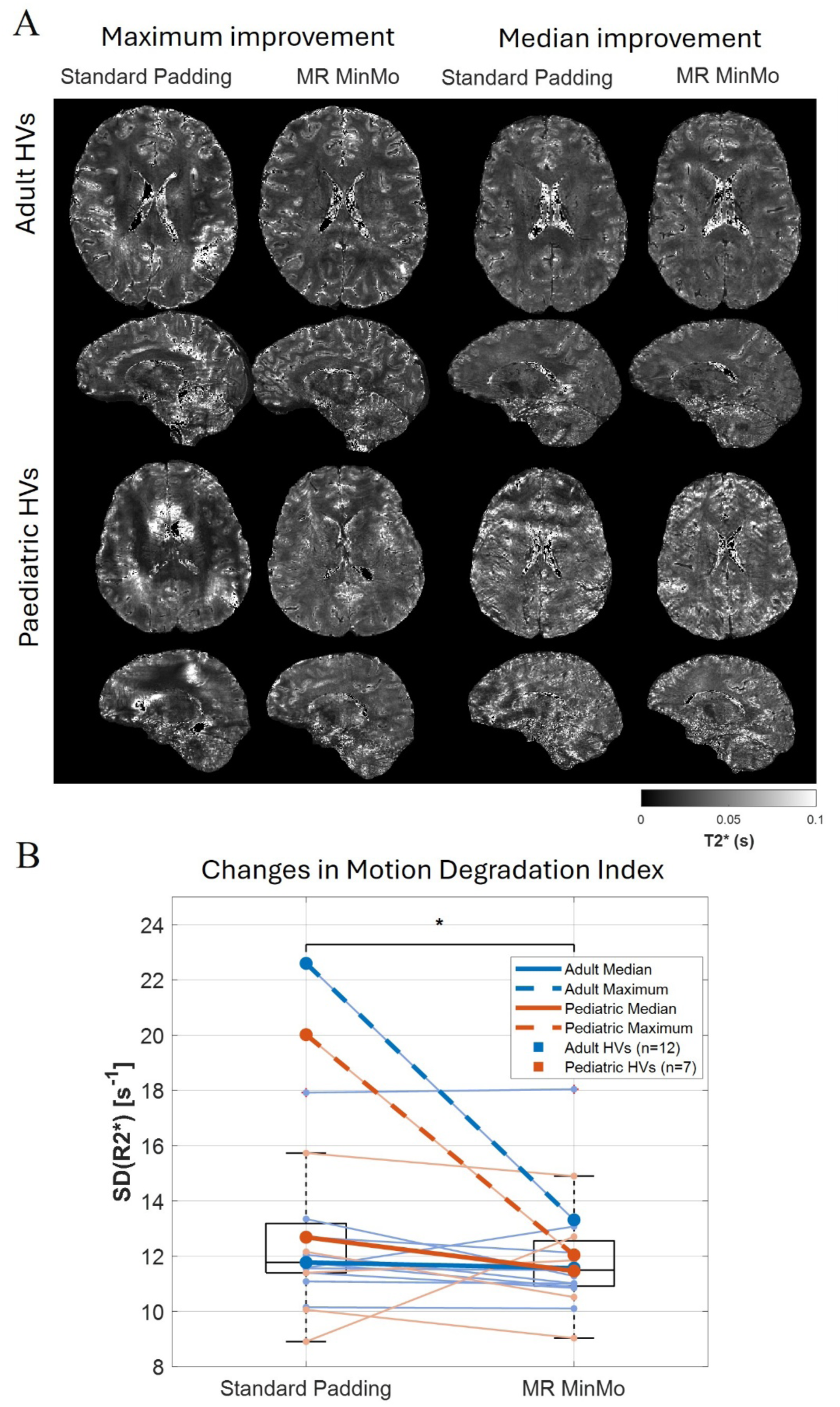
(A) Representative T2* maps (axial and sagittal views) shown for Standard Padding and MR-MinMo cases obtained by fitting across the ME-GRE echoes of the linearly sampled acquisition for the Adult HV and Paediatric HV with the maximum improvement and median improvement in SD(R2*) values. (B) Boxplot showing the motion degradation index SD(R2*) for standard padding and MR-MinMo conditions across adult (blue) and paediatric (red) participants. Individual subject trajectories are shown with light connecting lines, while representative cases with median (solid lines) and maximum (dashed lines) improvement are highlighted. Statistical significance is indicated by asterisk (paired t-test done for all HVs).

Representative participants, as shown in Figure 7(B)), benefited from a range of improvement, with adult HVs showing 2% (median) to 41% (maximum) SD(R2*) reduction, while paediatric HVs demonstrated 10% (median) to 40% (maximum) reduction. The significant maximum improvements in both groups show the ability for MR-MinMo to facilitate quantitative and qualitative motion suppression in select cases, while median improvements represent more general expected results across the cohort of volunteers.

## Discussion

We demonstrated that the MR-MinMo device was highly effective in reducing movement in the context of long-duration 7T scans. There was clear improvement in image quality both visually and quantitatively for scans of either ∼10 minutes or ∼20 minutes duration. Measures of motion estimated from retrospective correction showed significant reductions in maximum and average framewise displacement. The MR-MinMo was effective across participants in both compliant adult volunteers and younger paediatric volunteers. The younger group, who were also MRI naïve, tended to move more than older volunteers and therefore the MR-MinMo had the largest effect on image quality in this group. Quantitative R2* maps from the linear sampling protocol demonstrated a significant reduction in motion artefacts and increased precision, consistent with improvements in ME-GRE images. The 0.6mm isotropic resolution ME-GRE scans are typical of those used in quantitative multiparametric mapping studies^21,22^ at 7T, demonstrating MR-MinMo effectiveness in this context.

DISORDER sampling scans with 20-minute duration are deliberately motion sensitised to allow for motion estimation and subsequent correction. Retrospective correction significantly improved image quality, and we found an interaction between MR-MinMo use and retrospective correction suggesting synergistic effects. Alternative motion correction methods using navigators^23–25^ or tracking systems^1,26–28^ reliant on additional hardware/software may also benefit from reduced head pose variation to maintain data within a correctable regime.

Accurate coil sensitivity estimation is crucial for robust parallel imaging reconstruction^29^ but Participant motion affects receiver sensitivity profiles, particularly near small, dense receiver coil arrays, introducing potential reconstruction errors^6^. Movement can also affect B1+ field variability and while this is typically considered a minor contributor to reconstruction errors, minimising motion-related variance at the source can help reduce any residual variance. Suppressing motion with MR-MinMo helps reduce this error source, explaining the significant interaction between MR-MinMo and retrospective motion correction. However, we did not directly assess these factors.

Previous immobilization attempts include devices such as vacuum cushions, effective in various contexts^30–36^ but not routinely used despite availability, bite bars^37^ and individualised head moulds or thermoplastic masks^38^. Custom individualised foam inserts show mixed results with reports of limited efficacy^39,40^ and effectiveness. Bite bars and thermoplastic masks require custom manufacturing limiting deployment and integration into clinical workflows with additional issues of increased discomfort, feelings of claustrophobia and a sense of constriction, potentially exacerbating movement especially in paediatric use. This is problematic for long scans such as high-resolution whole brain structural MRI at 7T. We demonstrated encouraging results with clear improvements in adults and children using a device customisable in situ across a wide age range. In this context we have shown encouraging results with clear improvements in image quality in adults and children with a device customisable in situ to the individual across a wide age range. Due to space restrictions within the RF coil, subjects with larger head sizes may not be positioned comfortably with or without MR-MinMo.

Other motion mitigation strategies involve real-time tracking of head position using MRI acquisition adaptions (navigators), external devices such as optical cameras^41,42^, acoustic or ultrasound sensors^43–45^, inertial/mechanical devices^46,47^ or RF-sensor-based hardware^48^ showing improvements in motion-related artifacts. However practical limitations have restricted routine use^49^. Should these be overcome, these strategies can be employed, and are complementary to, improved head stabilisation.

### Limitations

Most adult HVs that were scanned were accustomed to 7T scanning; however, all paediatric HVs were scanned for the first time. While no formal survey was conducted due to ethics limitations, anecdotally most reported increased comfort with the MR-MinMo, consistent with reduced movement in very long scans estimated by DISORDER framework because reduced movement is only likely if volunteers were comfortable. Future systematic feedback would be needed to confirm this.

In all adult HVs except one, the MR-MinMo latch successfully closed for scanning in closed configuration. For one adult HV with larger head circumference, the Halo was removed and scanning proceeded in the open configuration. This volunteer (HV3) showed increased motion artefacts in the 20-minute scan with MR-MinMo compared to standard padding, however the DISORDER framework still increased image quality. Adult participants were also given the option to watch TV; however, some reported difficulty viewing the full screen extent while the MR-MinMo was in use due to standard mirror size and positioning limitations, which could be overcome with a custom mirror.

For visual comparison, we used native images without coregistration between conditions. While anatomy is not identical, this maintains fidelity regarding PE directions differentially sensitive to motion artefacts while avoiding potential interpolation artefacts such as partial volume effects or overshooting/ringing artefacts^50^.

Some residual artefacts potentially pertaining to Gibb’s ringing may remain in some HVs but were consistent across conditions and could be removed by k-space filtering techniques. We avoided filtering of any kind for clearest visualisation and measurement of any differences between standard and MR-MinMo stabilisation.

We evaluated intrascan motion, but many quantitative protocols (e.g. MPM^21,51^) consist of acquiring several images with different parameters (e.g. flip angle). Although not evaluated here, reductions in interscan movement might improve quantitative mapping by reducing position-dependent transmit and receive field effects^52^ – this should be explored in future studies.

The MR-MinMo use was randomised across all HVs; 9 of 19 HVs were scanned with the MR-MinMo first, 10 with MR-MinMo second, ensuring that any increase in motion prevalence with time in the scanner^53^ should not have affected results.

No direct comparison was made to other head stabilisation methods such as vacuum cushions, so relative efficacy cannot be unambiguously determined. None-the-less comparison to current practice showed clear advantages in image quality from improved head stabilisation with and without retrospective correction.

## Conclusion

The MR-MinMo was found to be an effective solution for reducing head motion and increasing image quality in high resolution 7T scans in adults and particularly children. The device is compatible with motion correction methods and was shown to improve the performance of the DISORDER retrospective motion correction method.

## Data Availability Statement

Representative T2* quantitative maps and analysis code are available at [URL will be inserted upon acceptance]. The MR-MinMo device design specifications are available from the corresponding author upon reasonable request. Source code for the DISORDER reconstruction and motion analysis is available at [URL available upon acceptance]. Raw imaging data are available from the corresponding author upon reasonable request, subject to ethical approval and data sharing agreements.

